# Operational Strategies among Infectious Disease Clinical Trial Sites During Pandemics: A Scoping Review

**DOI:** 10.1101/2025.10.15.25338123

**Authors:** KL Trigg, A Zhang, AW Wu

## Abstract

**Purpose:** To examine documented strategies, challenges, and lessons related to clinical trial site operations during pandemics.

**Methods:** This review focused on seven major pandemics: COVID-19, HIV/AIDS, Ebola, SARS, MERS, H1N1, and Zika. Searches were conducted using Pubmed and Embase between September 2024 and February 2025, yielding 7,572 unique records. After dual screening, 47 articles met inclusion criteria, with an additional 8 identified through citation searching, for a total of 55. Non-English articles were translated using Google Translate. Data were extracted and thematically analyzed through iterative familiarization, keyword identification, coding, and consensus discussion.

**Results:** Of the 55 included articles, most originated from the European Region (61%) and the Americas (52%), with additional representation from Africa (19%), the Western Pacific (8%), Eastern Mediterranean (5%), and Southeast Asia (5%). Percentages exceed 100% because several studies reported across multiple regions. Study designs were predominantly descriptive (46%) and observational (30%), with fewer randomized controlled trials (22%) and cohort studies (2%). COVID-19 accounted for the majority of articles (56%), followed by HIV/AIDS (26%), Ebola (10%), H1N1 (4%), and Zika (3%); no eligible studies addressed SARS or MERS.

Operational themes included policy (73%), resources (55%), networks (53%), infrastructure (47%), technology (45%), study design (31%), and communication (22%). Publications peaked during pandemic years, reflecting intensified research activity. Reported challenges included protocol–practice misalignment, fragmented infrastructure, regulatory bottlenecks, and under-resourced sites. Effective practices included early site engagement in protocol development, streamlined ethics and contracting processes, investment in digital and decentralized infrastructure, and cross-trained staff. Gaps included limited use of adaptive trial designs and insufficient cross-national collaboration.

**Conclusions:** Clinical trial sites implemented systemic operational changes to sustain research during pandemics. Key lessons include the need to strengthen infrastructure, workforce capacity, and institutional adaptability, and the need to address persistent global inequities. Institutionalizing early site engagement, adaptive designs, and equitable collaboration will be essential to enable timely and resilient clinical research responses in future pandemics.

## Introduction

Despite the critical role of clinical trials in advancing medicine, their conduct is often characterized by conservative practices, operational inefficiencies, and protracted timelines. Before the COVID-19 pandemic, the most commonly cited reasons for trial discontinuation were overestimated prevalence of the condition, misjudged interest in the intervention, under-recognized administrative or participant burden, and inadequate funding(1). During pandemics, clinical trial sites are faced with additional challenges, especially concerning trial data integrity(2,3).

Coupling the new challenges associated with pandemics with the typical approval timeline for a new drug of 10 years(4–6), and the small number of approved antibiotics (5.29% of the total approved) and antivirals (5.96%) since 2000(7), there is an unparalleled need to improve clinical trial operations. Recently, drug approval priorities had shifted to approving anticancer drugs, with nearly half of new drug submissions during COVID-19 involving anticancer agents(7). Yet, research funding and activity in infectious diseases do not align with the regulatory approvals of new medicines.

There was an urgent need during the COVID-19 pandemic to develop novel antiviral agents and to use of accelerated approval pathways, sometimes referred to as fast-track(3,7–9). Accelerating the approval phase of novel agents is essential during an outbreak. However, even when this is successful the new pathways may not be sustained once an outbreak is over. However, the high prevalence of infectious diseases, coupled with their disease burden and socioeconomic costs, provides ample evidence for regulators to promote sustained efficiency beyond pandemics.

Moreover, clinical research sites, even those located at Academic Medical Centers (AMCs) which represent the majority of sites globally(10,11), are frequently excluded from decision-making processes and the design of industry clinical trials(12–14). This lack of engagement perpetuates arcane models and processes(15). Examining real-world examples of innovations at clinical trial sites during pandemics can provide valuable lessons(16), while also highlighting that economic constraints, infrastructure limitations, and fragmented operational processes are often under-addressed in the literature on clinical trial operations.

There is limited research on the challenges faced by infectious disease clinical trial sites during outbreaks, and how they adapt operational workflows and processes during pandemics. This scoping review aims to identify and synthesize the strategies developed by infectious disease clinical trial sites in response to six pandemics: COVID-19, HIV/AIDS, Ebola, SARS, MERS, H1N1, and Zika.

This scoping review drew evidence from the published literature to offer practical, context-specific insights that can help sites prepare for future outbreaks. The objectives of this review were to identify the challenges, solutions, and opportunities associated with conducting infectious disease clinical trials during pandemics.

## Methods Design

### Patient and Public Involvement

Patients were not involved in this research.

### Search Strategy

A scoping review was conducted to identify strategies implemented by infectious disease clinical trial sites in response to operational challenges that arise during pandemics. Challenges were identified using an inductive approach, aiming to capture existing knowledge and allow new themes and patterns to emerge from the included articles. Adaptations or solutions to identified challenges were classified as interventions. Protocols and randomized controlled trials were included only if they described operational interventions. Studies, commentaries, opinions, and editorials were included based on the general criteria identified in the Population, Intervention, Comparator, Outcome, Time and Setting (PICOTS) framework (Table 1).

**Table 1.** Population, Intervention, Comparator, Outcome, Time and Setting (PICOTS) framework.

| PICOTS | Inclusion criteria | Exclusion criteria |
| --- | --- | --- |
| Populations | Infectious disease clinical trial units or directly related operational units with modifications to existing processes or methods in clinical trial conduct during pandemics, epidemics and outbreaks | None |
| Intervention | Technology<br>Process/workflow/pathway adaptation<br>Decentralization<br>Institutional adaptations<br>Staffing<br>Accelerations | Descriptive studies or studies outside Infectious diseases |
| Comparators | N/A (status quo) | N/A |
| Outcomes | Good Clinical Practice<br>Patient burden<br>Site process improvement<br>Simplifying regulatory processes<br>Inequity / Access<br>Timelines<br>Infrastructure | Does not discuss or describe the listed outcomes of interest<br>Studies not describing intervention or adaption during infectious emergency |
| Timing | Studies completed since 2000 | Studies completed prior to 2000 |
| Setting | All care settings (academic medical center, private practice)<br>Global | Industry only management<br>Clinical trial reports/protocols without descriptions of adaptations/novel methodologies |

PICOTS = populations, interventions, comparators, outcomes, timing, setting

To ensure methodologic rigor and transparency, Joanna Briggs Institute’s (JBI) scoping review methodology(17) was followed and the Preferred Reporting Items for Systematic reviews and Meta-Analyses extension for Scoping Reviews (PRISMA-ScR)(18) (Table 2) was utilized to report findings of this scoping review. This review was guided by the following research question: *What strategies, challenges, and lessons have been documented in the literature regarding clinical trial site operations during pandemics?*

Searches were conducted in PubMed and Embase from September 2024 through February 2025 using the search criteria detailed in Table 3. No restrictions were applied by language nor publication date. Search strings combined Medical Subject Headings (MeSH) and Title/Abstract keywords to capture article type, clinical trial site operations, and the six pandemics of interest for this review.

**Table 3.** Search Criteria in PubMed.

| Search # | String | Notes | Results |
| --- | --- | --- | --- |
| 1 | "clinical trial" [mesh] OR clinical trial[tiab] OR "clinical trial"[pt] OR "clinical research" [mesh] OR clinical research[tiab] OR "clinical research"[pt] OR "clinical study"[Mesh] OR clinical study[tiab] OR "clinical study"[pt] OR intervention study[tiab] OR "intervention study"[Mesh] OR "intervention study"[pt] OR "evaluation study"[Mesh] OR "evaluation study"[tiab] OR "evaluation study"[pt] OR "randomized controlled trial"[Mesh] OR randomized controlled trial[tiab] OR "randomized clinical trial"[pt] "adaptive trial"[Mesh] OR adaptive trial[tiab] OR "adaptive trial"[pt] OR "adaptive study"[pt] OR "adaptive study"[Mesh] OR adaptive study[tiab] OR "protocol, clinical"[Mesh] OR protocol, clinical[tiab] OR "clinical protocol"[Mesh] OR treatment protocol[tiab] OR "treatment protocol"[Mesh] OR protocols, treatment[tiab] OR "protocols, treatment"[Mesh] OR clinical research protocol[tiab] OR "Research Protocols, Clinical" [Mesh] OR "Protocols, Clinical Research" [Mesh] | Study type string | 13,531 |
| 2 | ("site"[Title/Abstract] OR "clinical trial site" [Title/Abstract] OR "clinical research site" [Title/Abstract] OR "clinical research clinic"[Title/Abstract] OR "academic medical center"[Title/Abstract] OR "university"[Title/Abstract] OR "network"[Title/Abstract] OR "clinical trial* network"[Title/Abstract] OR "consortium"[Title/Abstract] OR "initiative"[Title/Abstract]) | Clinical trial sites | 2,188,150 |
| 3 | (1 AND 2) |  | 1,200 |
| 4 | "COVID-19"[Mesh] OR "COVID-19"[tiab] OR "SARS-CoV-2"[tiab] OR "2019 Novel Coronavirus"[tiab] OR Pandemics[Mesh] OR pandemics[tiab] OR pandemic[tiab] | COVID-19 | 431,644 |
| 5 | "severe acute respiratory syndrome"[tiab] OR "Severe Acute Respiratory Syndrome"[Mesh] | SARS (Not COVID-19) | 48,649 |
| 6 | ("Hemorrhagic Fever, Ebola"[Mesh]) OR "Ebolavirus"[Mesh] OR ebola[tiab] "SARS COV"[tiab] | Ebola | 585 |
| 7 | "Middle East Respiratory Syndrome Coronavirus"[Mesh] OR "Middle East Respiratory Syndrome"[tiab] OR MERS[tiab] | MERS | 9,213 |
| 8 | "Influenza A Virus, H1N1 Subtype"[Mesh] OR H1N1[tiab] OR "swine flu"[Mesh] OR "influenza"[Mesh] | H1N1 | 24,102 |
| 9 | "Zika" [Mesh] OR Zika[tiab] | Zika | 11,691 |
| 10 | "Human Immunodeficiency Virus"[tiab] OR HIV[tiab] OR AIDS[tiab] | HIV | 478,359 * |
| 11 | (4 OR 5 OR 6 OR 7 OR 8 OR 9 OR 10) |  | 1,004,416 |
| 12 | 3 and 11 |  | 6,560 |

The results were uploaded to DistillerSR (DistillerSR. Version 2. DistillerSR Inc.; 2025. Accessed Sept 2024-Feb 2025.) and de-duplicated for screening. Titles and abstracts were screened by one reviewer against eligibly criteria, with a secondary reviewer independently assessing 10% of excluded records. Discrepancies regarding eligibility were resolved through discussion.

Once articles were determined in the initial search, forward and backward citation (snowballing or citation chasing) searching was performed on the included articles in PICO Portal (PICO Portal. Version 3.0.2025.0225. PICO Portal; 2025. Accessed Nov 2024-Feb 2025), identifying an additional 2,539 articles. These were uploaded and de-duplicated in DistillerSR and screened using the same process described above. Adjudication was again conducted on a 10% sample of excluded records and full texts, with discrepancies resolved through discussion.

Grey literature was not systematically searched, given the limited availability of reliable sources describing operational changes and site-level experiences during pandemics.

### Data Extraction and Synthesis

#### Thematic assignment

Themes were identified through a systematic process of data familiarization, keyword selection, coding, and iterative discussion. Articles were first categorized by content to generate initial keywords and themes, then reviewed in tabular format to identify recurring patterns. Keywords were derived inductively from the data and refined through consensus among the research team, leading to final thematic assignments.

#### Evidence Table Generation

An evidence table was developed (Table 4) to extract the following variables: first author, year of publication, country(ies) of all affiliated authors, disease(s), study type, keywords, and themes. Given the broad range of study types and time periods included, a customized extraction table was designed to capture the main theme(s) and relevant subcategories. A second reviewer conducted a confirmatory review, and disagreements were resolved by consensus. Inclusion of all affiliated author countries was intended to highlight the global scope of collaborative research during infectious disease emergencies.

**Table 4.**
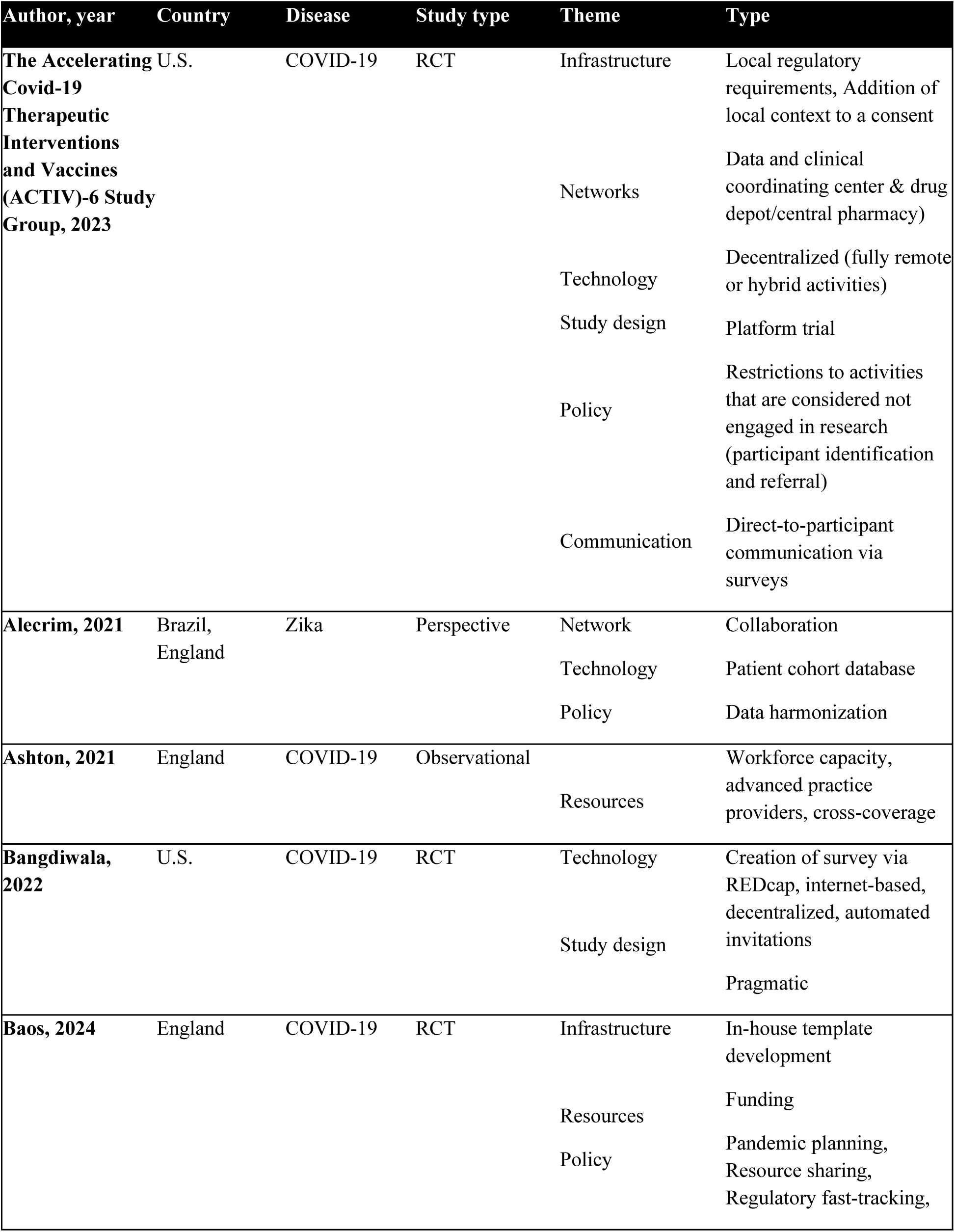
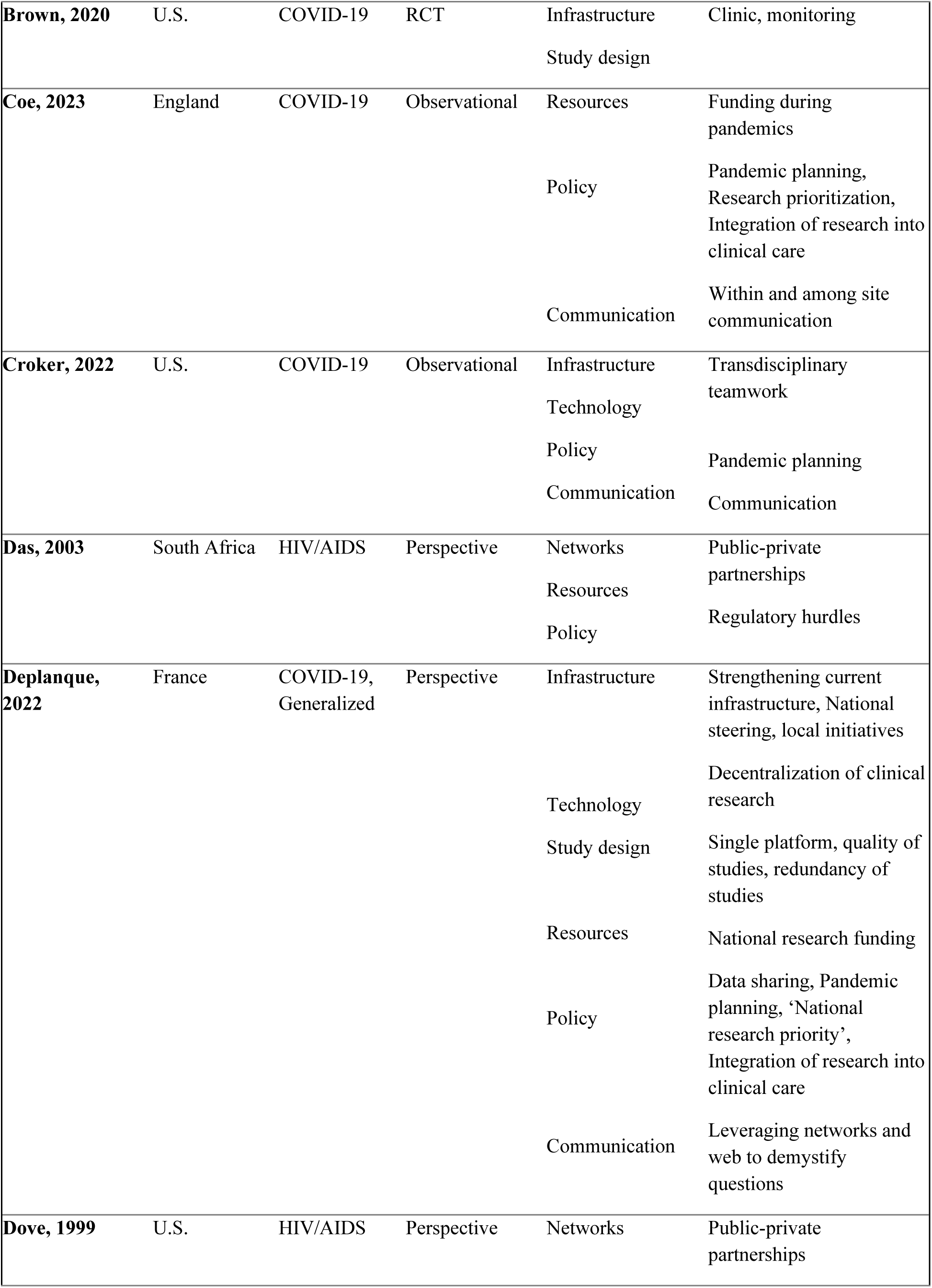
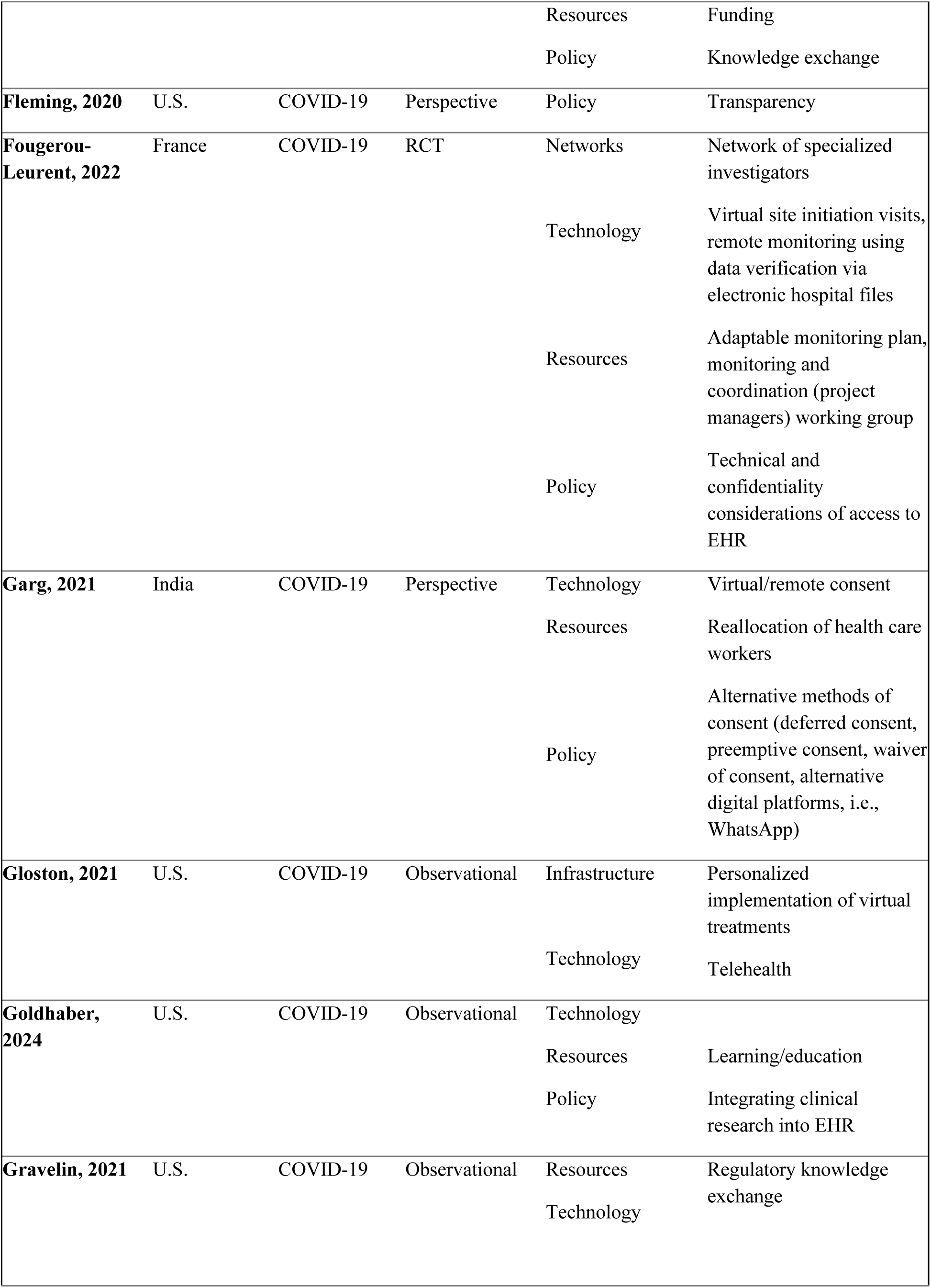
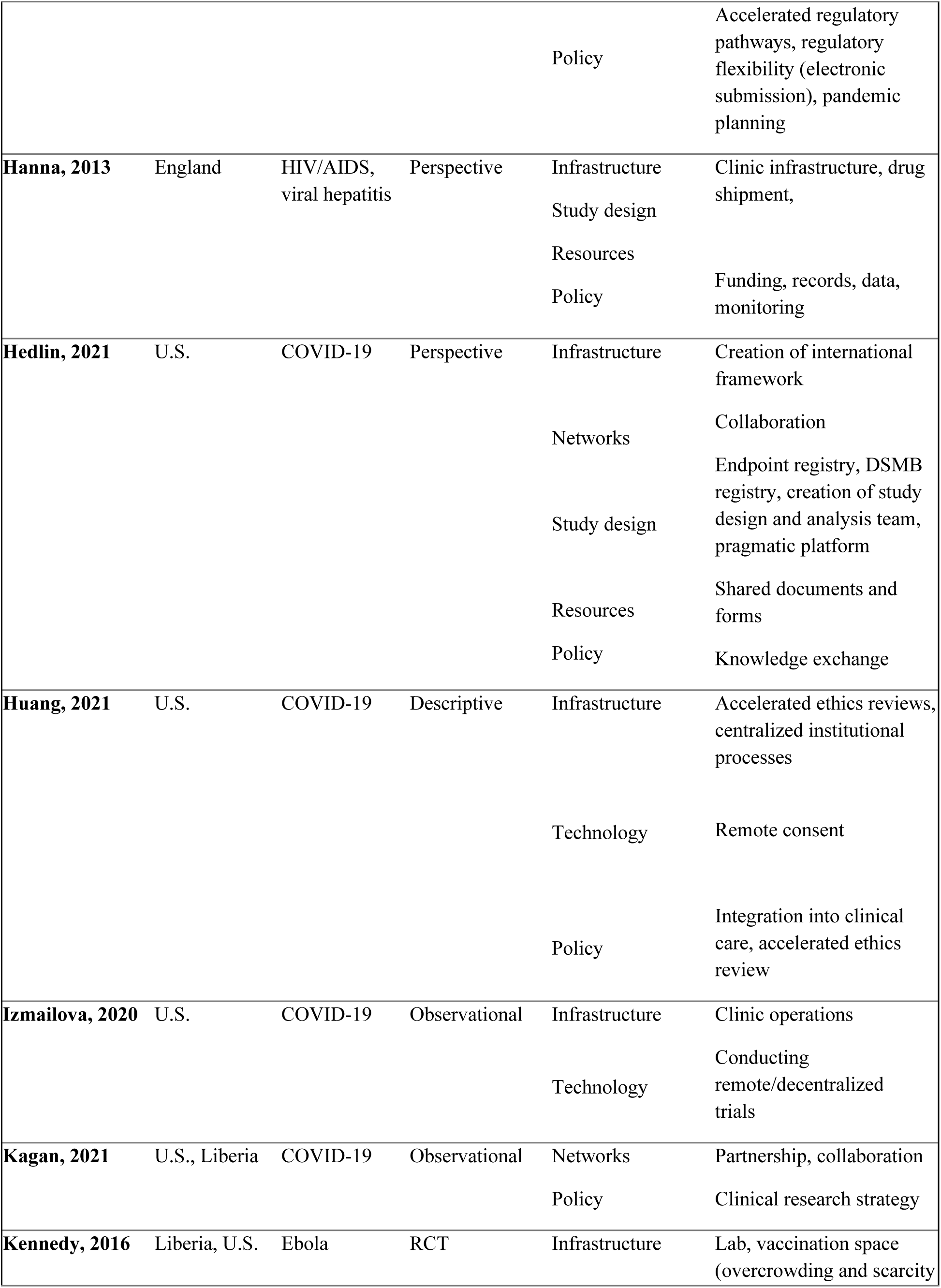
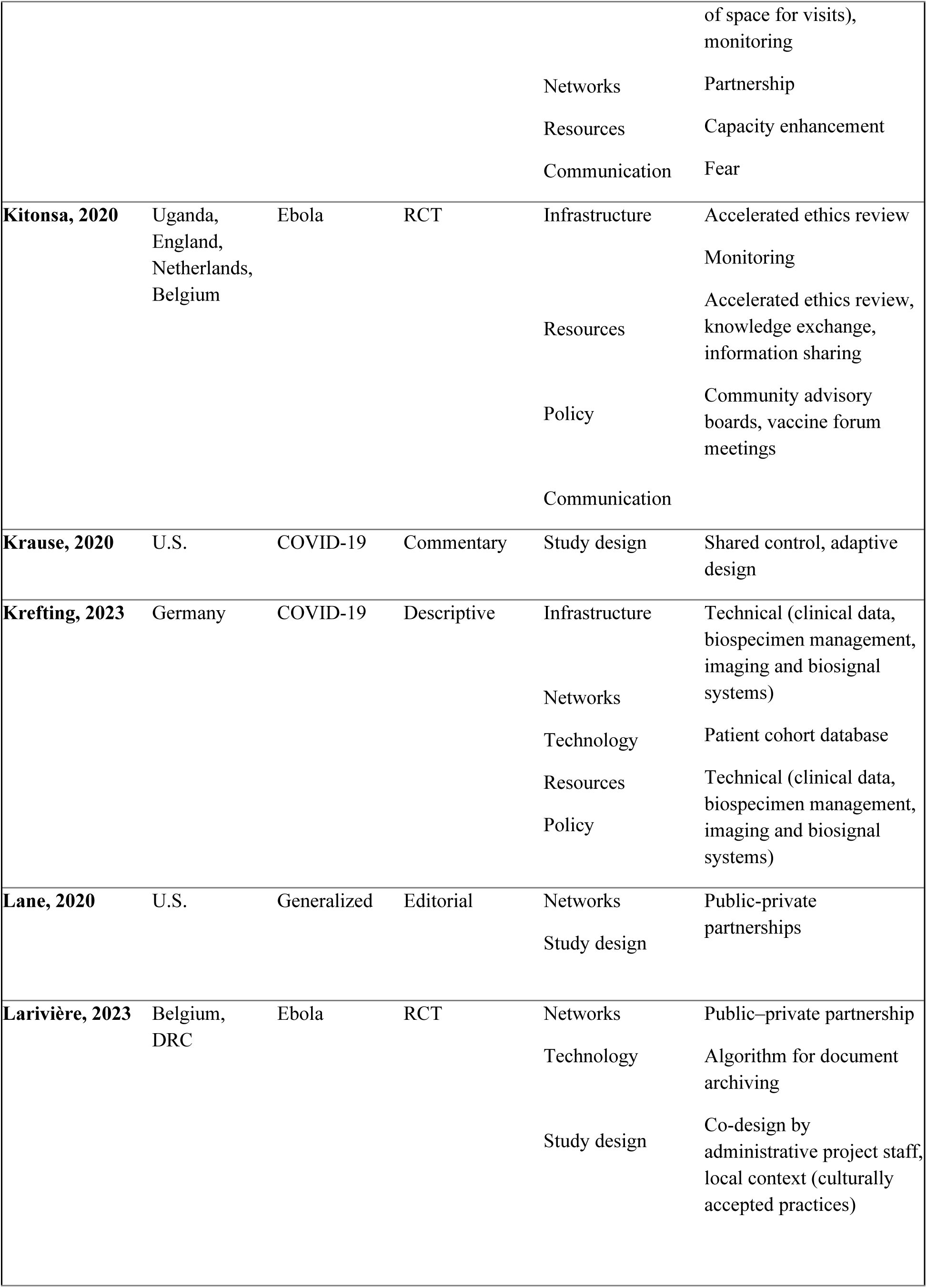
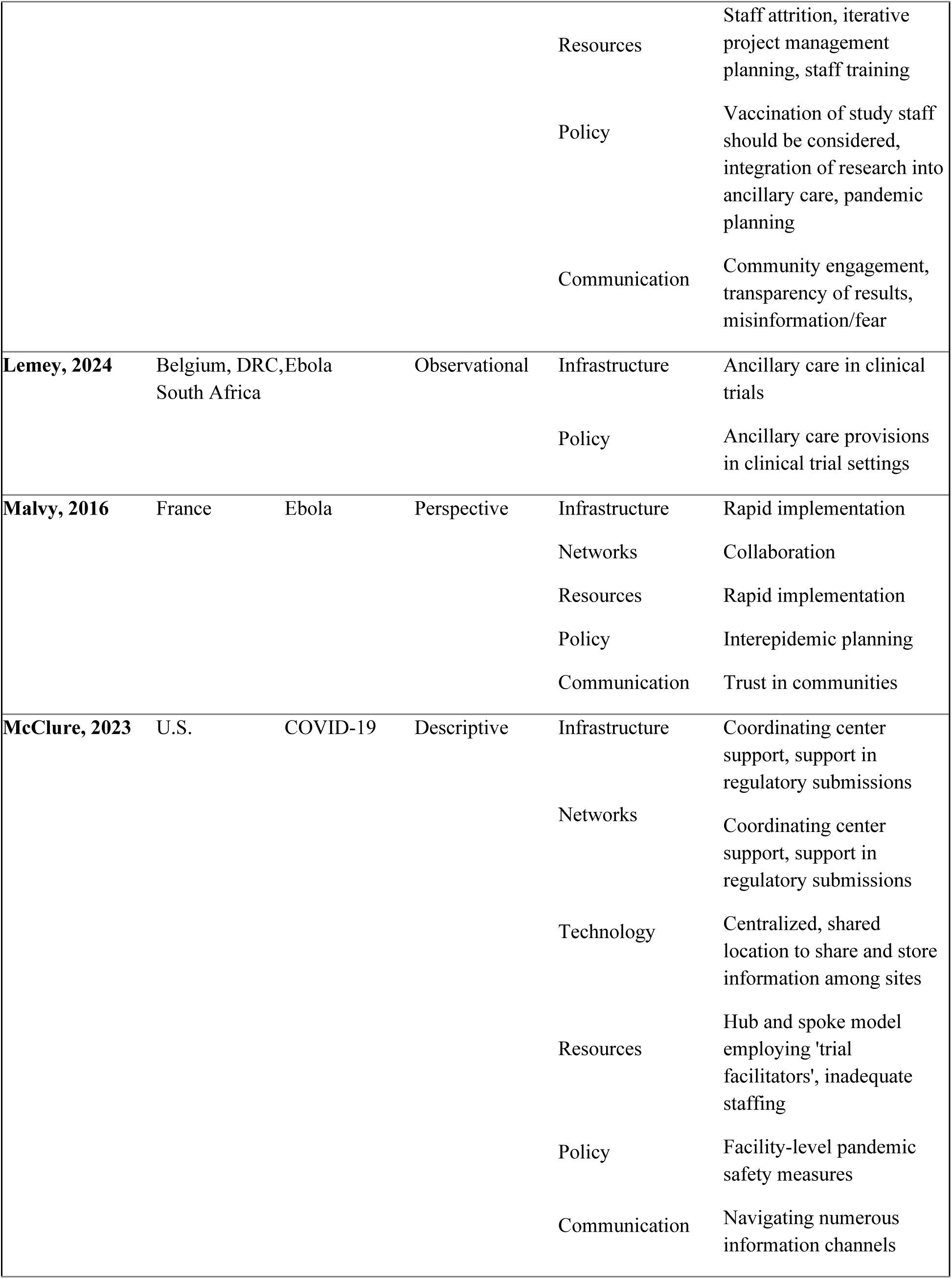
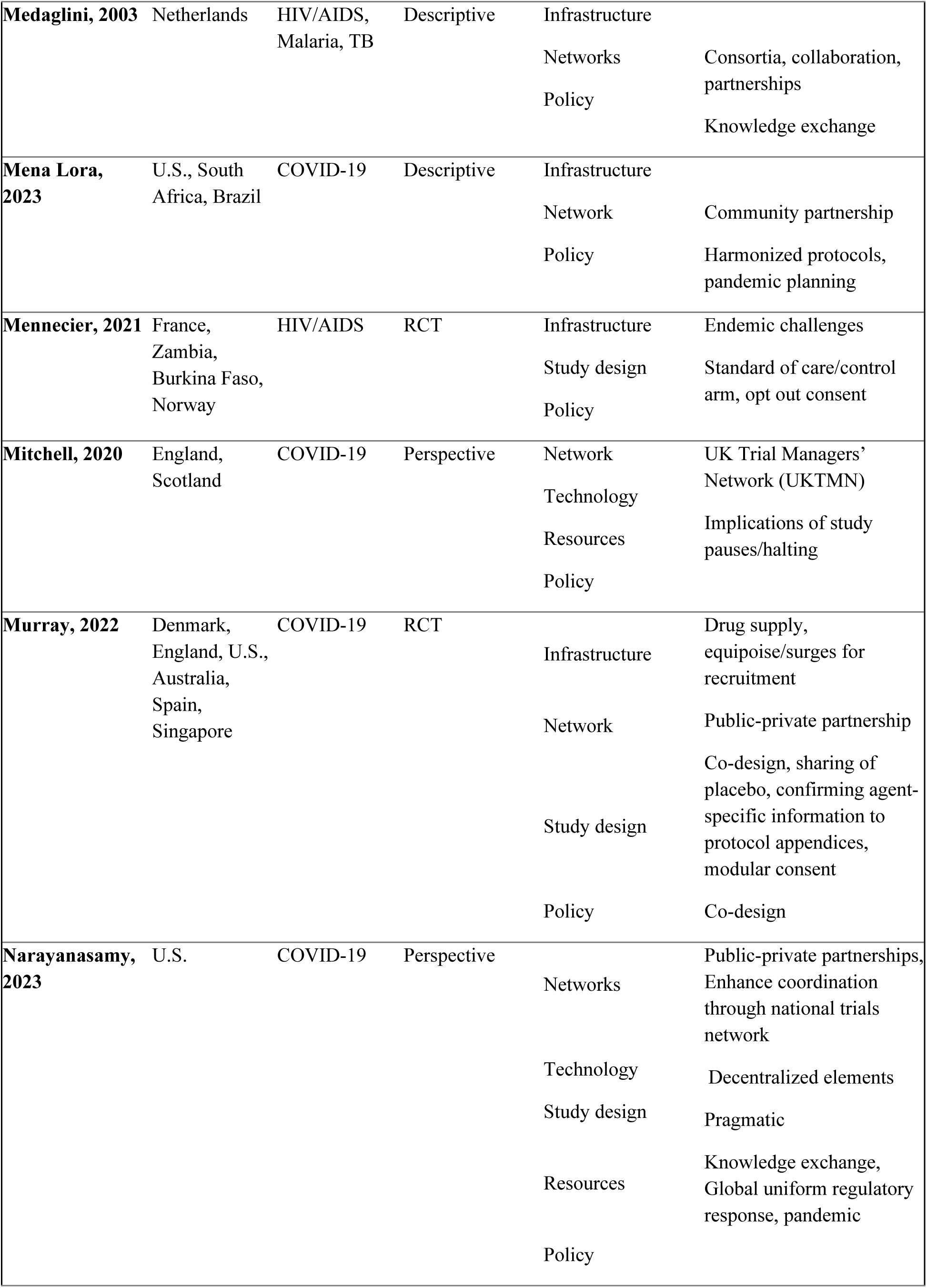
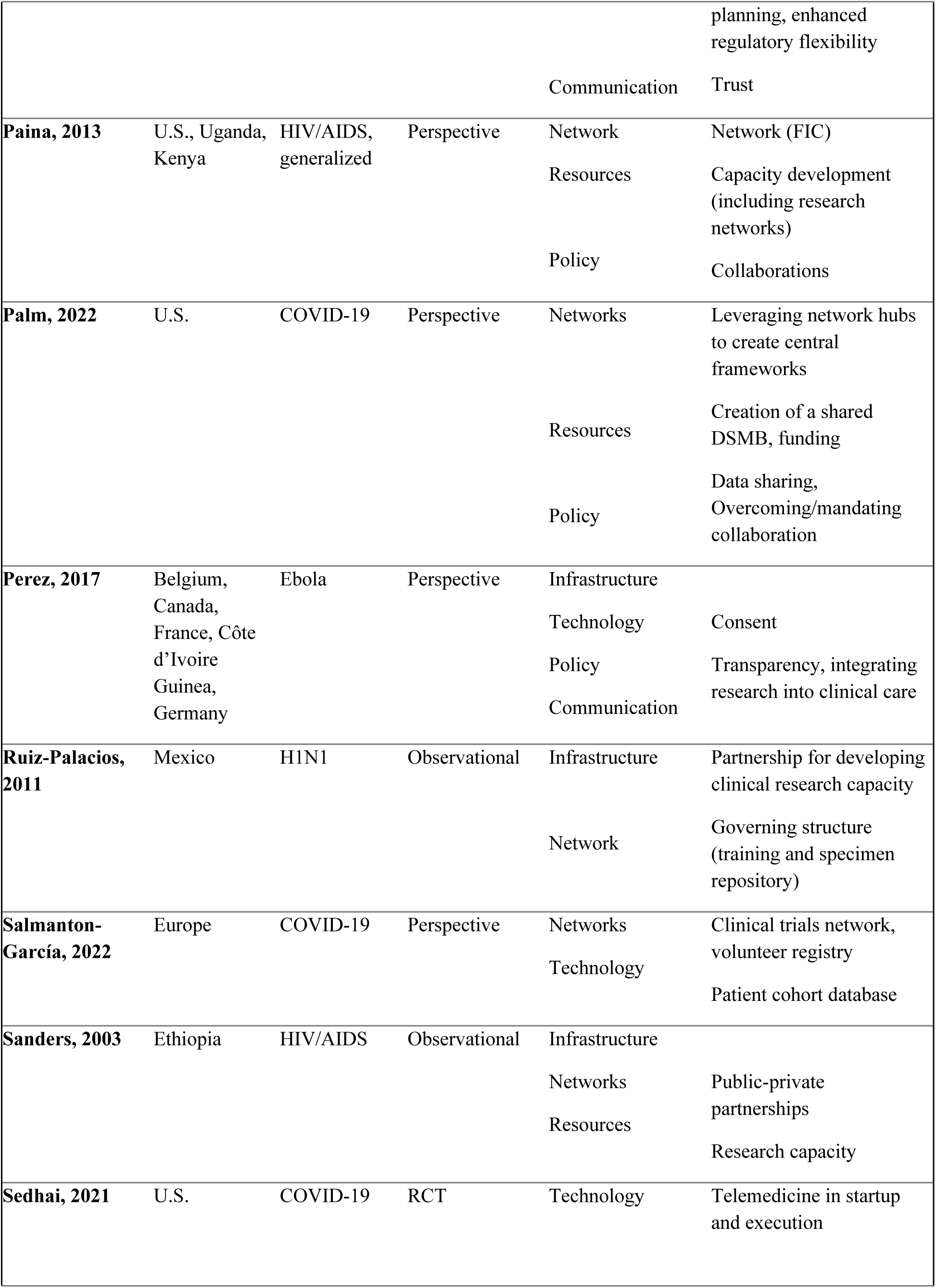
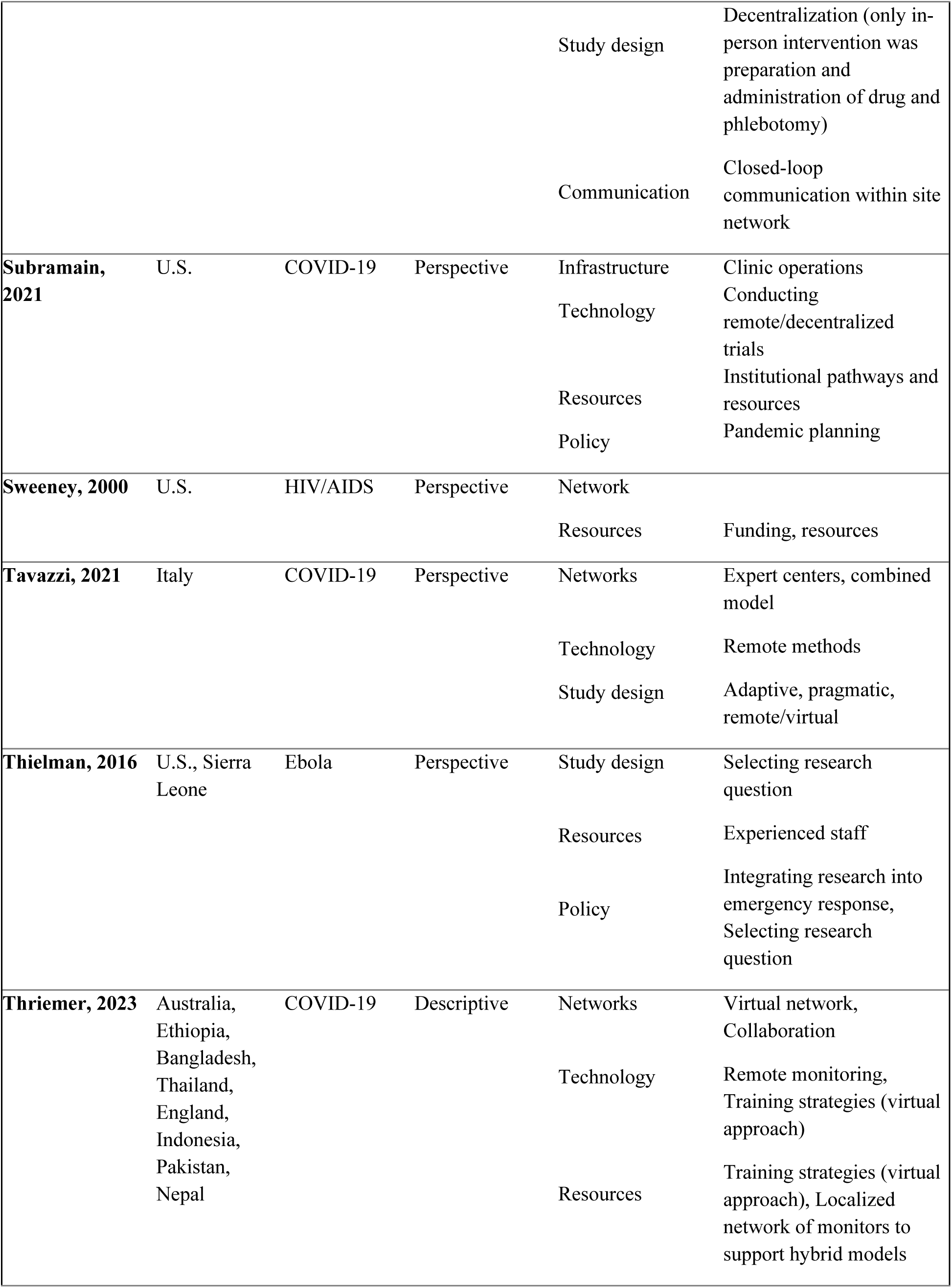
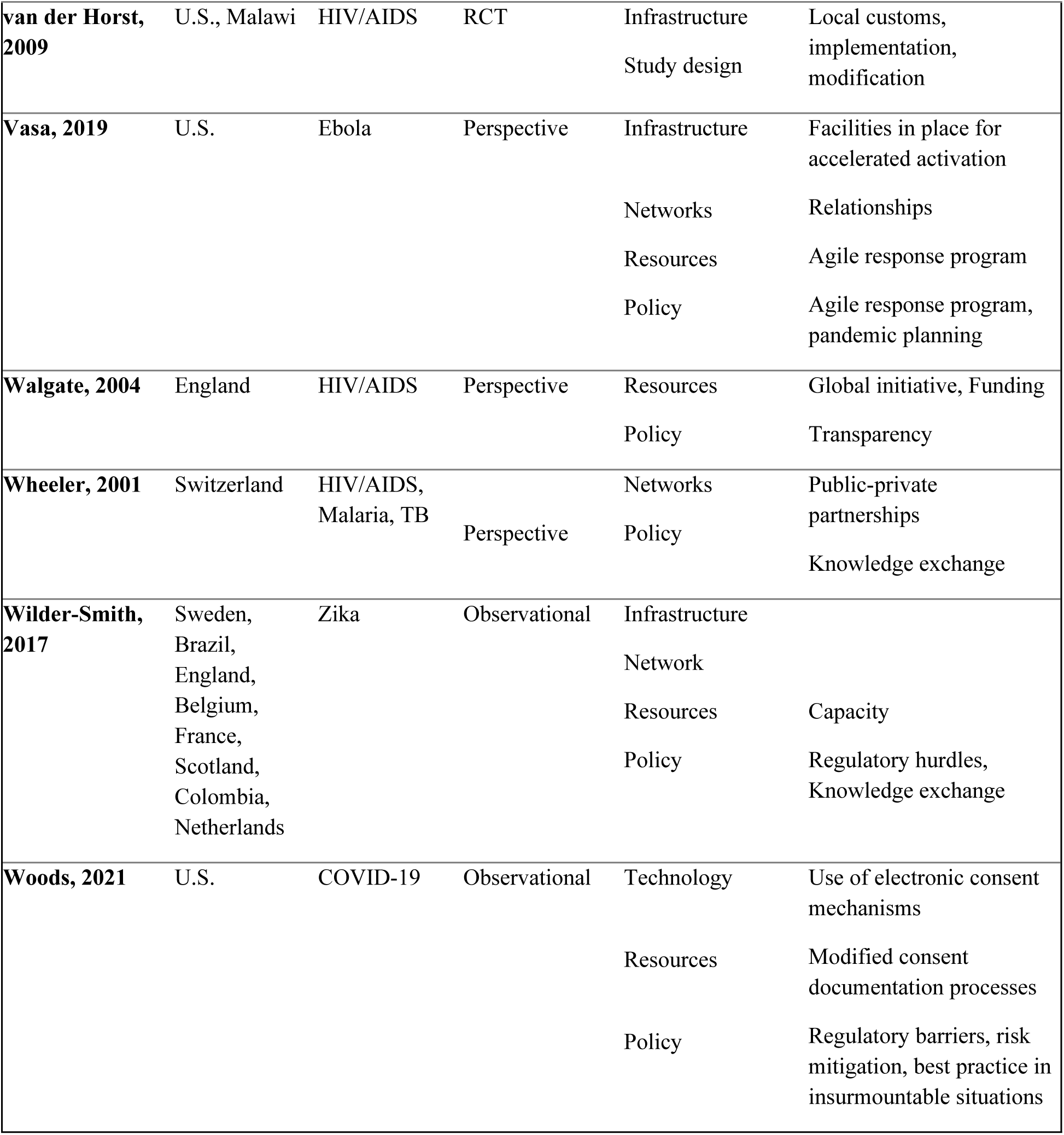
Evidence Table.

## Results

A total of 8,171 records were identified from PubMed and Embase. After deduplication, 7,572 unique records remained; 6,931 were excluded and 641 underwent secondary screening, yielding 47 eligible articles. Forward and backward citation screening of these 47 papers identified an additional 2,539 records; after deduplication, 8 more publications were included, for a final sample of 55 (Figure 1).

**Figure 1.**
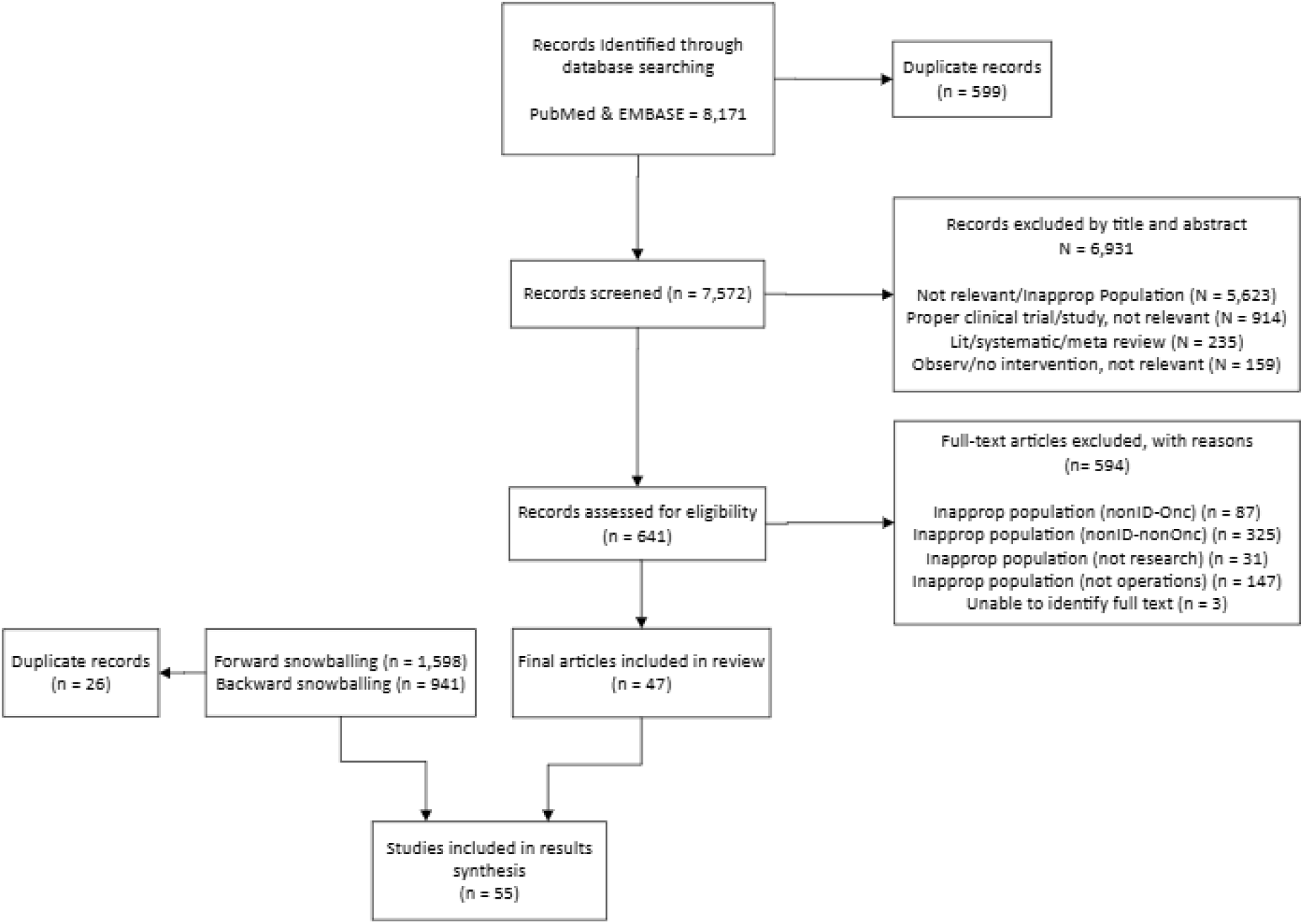
PRISMA Diagram for scoping review study selection.

### Types of included articles

Publications spanned 1999–2024, with sharp increases during pandemic years. COVID-19 accounted for 66% of all articles. Perspectives (editorials, commentaries, interviews, and opinion pieces) comprised 60% of the sample, while 40% were original research. Of these, randomized controlled trials represented 22% and observational designs 18% (including descriptive analyses, implementation studies, and published abstracts). Sixty percent (n=33) were peer-reviewed studies.

### Countries and regions

To assess geographical distribution, all author affiliations were collated and tabulated. In cases of multi-country authorship, all countries listed were included. For consistency and comparability, geographic representation was classified according to WHO regions (Figure 2).

**Figure 2.**
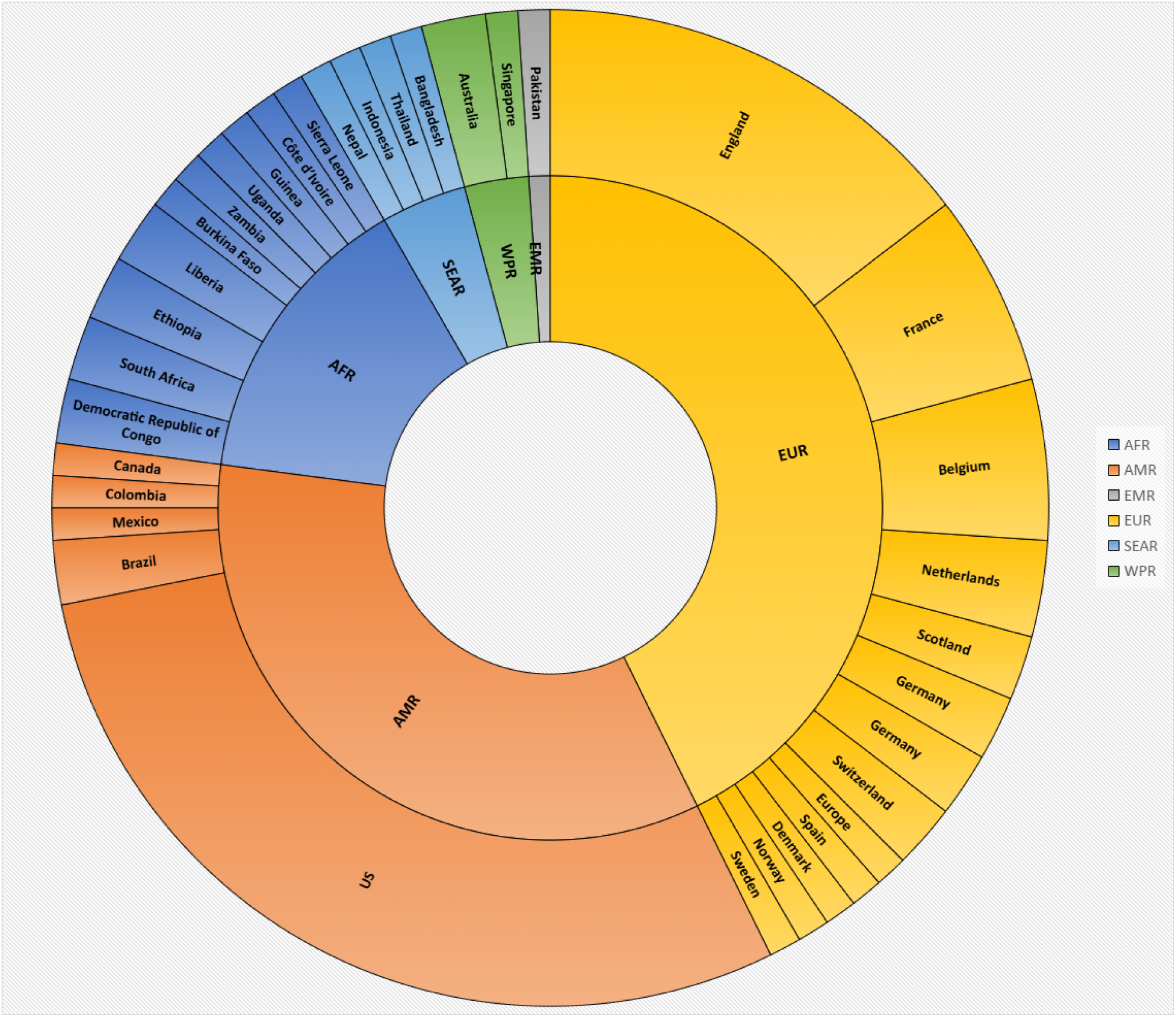
Geographical distribution of studies. Sunburst chart showing the hierarchical distribution of records identified by region and country. Segment size corresponds to the relative number of published records within each geographic level.

When examining the data by disease, the majority described responses to COVID-19 (32, 58%), followed by HIV/AIDS (11, 20%), Ebola (7, 13%), H1N1 (1, 1.8%), and Zika (2, 3.6%).

There were two articles that described generalizable reflections on pandemics without identifying a particular disease (2, 3.6%). There were several HIV/AIDS articles that described non-outbreak infectious diseases such as tuberculosis (2, 4%), malaria (2, 4%), and hepatitis (1, 2%). Articles could be classified under multiple pandemics. There were no articles on SARS or MERS found in this review.

### Distribution across domains

Articles were categorized into seven domains of trial operations: policy, resources, networks, infrastructure, technology, study design, and communication. Many articles were classified into multiple domains.

Of the 55 articles, policy was most frequently discussed (40, 73%), followed by resources (30, 55%), networks (29, 53%), infrastructure (26, 47%), and technology (25, 45%) (Figure 3).

**Figure 3.**
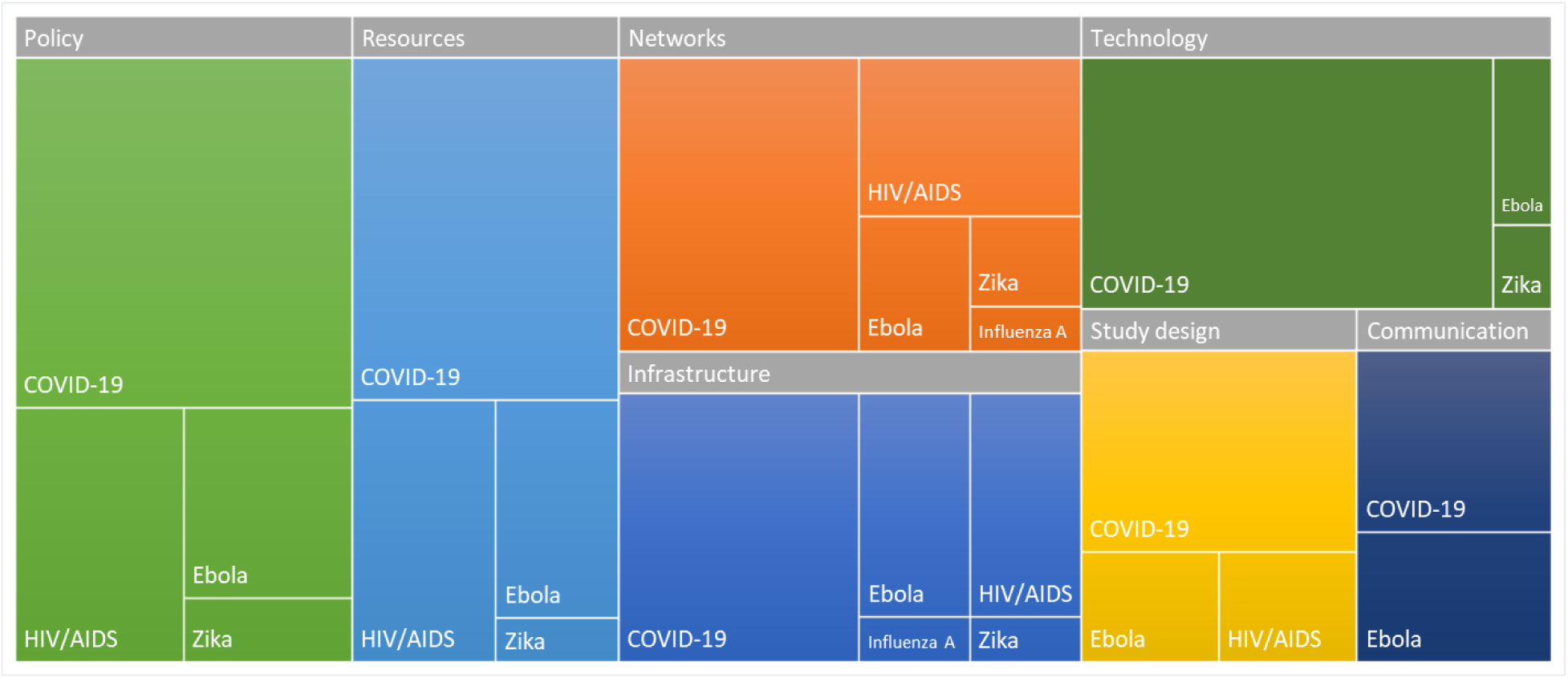
Treemap of themes broken down by disease (count of articles). The size of each rectangle corresponds to the proportion of each theme, allowing for visual comparison of relative contributions across themes.

Fewer addressed study design (17, 31%) and communication (12, 22%). Cross-classification was common, reflecting the interconnectedness of trial operations.

### Trends over time

Publication volume rose sharply during defined pandemic years (1999, 2003, 2013–2017, 2020), aligning with major outbreaks captured in this review. These surges reflected not only increases in the number of articles but also expansion in thematic coverage, with more domains of clinical trial operations addressed during outbreak periods compared to non-pandemic years (Figure 4).

**Figure 4.**
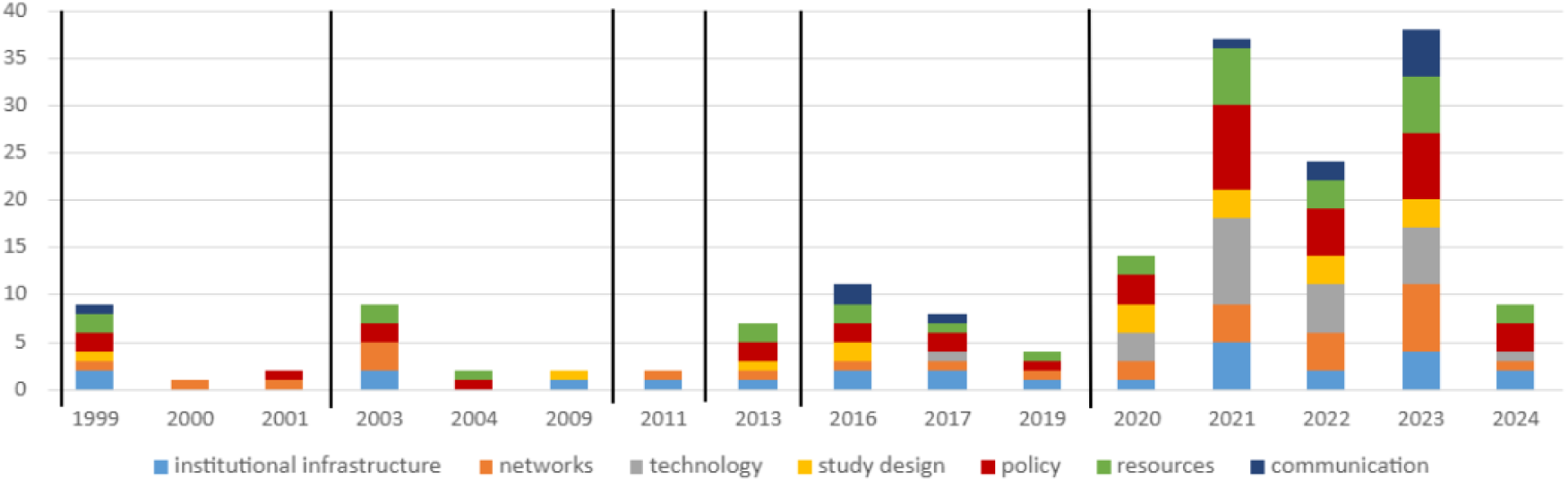
Themes identified over time with pandemics demarcated as vertical events. Vertical demarcation lines indicate the onset of the seven pandemics (HIV/AIDS pre-1999, SARS in 2002, H1N1 in 2009, MERS in 2012, Ebola in 2014, Zika in 2016, and COVID-19 at the end of 2019), marking key time points for contextualizing shifts in the volume and proportion of themes across time.

### Themes identified

Seven categories of themes were identified: policy, resources, networks, infrastructure, technology, study design, and communication. Most adaptations focused on establishing policies, leveraging of networks and partnerships, or urged improvements in infrastructure and need for collaborative approaches.

#### Policy

Robust policies for data sharing, governance, and monitoring were deemed essential(19–23). In addition, there is a need for better preparedness planning that includes clinical research(23–32), extending the concept of integrating clinical research with medical care as part of an outbreak response.

Ensuring data quality during an outbreak can be challenging for clinical trial sites(33,34), Articles described adapted monitoring plans, DSMBs, a COVID-19 dedicated design and analysis team(35), shared repositories of trial documents, a pragmatic platform protocol utilized during COVID-19 (35) and common data collection tools(33,35–39).

Barriers included institutional culture, conflict over data ownership and publication priority, and underestimation of the effort required for harmonization(36,40). The competitive structure of academic medicine can be at odds with the need for collaboration in a pandemic.

Integrating clinical research into clinical care is a topic mentioned across articles. Mechanisms included leveraging the use of workflows within the EHR. Clinical Research as a Care Option (CRAACO) embeds research into EHR systems and was widely praised as strengthening preparedness and recruitment capacity(41,42). The most significant factor on recruitment was integrating research recruitment into clinical care. To enable integration, sites need to achieve timely and sufficient participant recruitment, and there is an overwhelming need to prioritize research during future pandemics(23–32).

#### Resources

Funding and staffing shortages were recurring barriers(43–46). Articles noted gaps in personnel support(33,47), capacity building(24,31,48,49), and prioritization of research within care settings(42,46), and may extend to *<u>Infrastructure</u>* support within clinics, labs, hospitals, and administrative spaces to sustain clinical trial operations(47,49).

Clinical trials in resource-constrained settings face multiple barriers, including limited infrastructure, regulatory challenges, and difficulties in rapidly enrolling participants during outbreaks; these are compounded by the unpredictable and urgent needs that arise in pandemics(50)^49^. Additionally, the unique characteristics of each outbreak can affect the feasibility and design of clinical trials, especially related to prioritizing partnerships (49).

Resource limitations were compounded in low-resource settings(20,27,34,36,42–46,51), though lessons from these contexts were seen as transferable from lower to higher income settings. Redeployment and cross-coverage of staff emerged as critical strategies(52–54). Several frameworks, including adapted management models, were proposed to align strategy with operations and strengthen site capacity(55).

#### Network

Publications highlighted centralized coordination(23,56), statistical and data management centers(36), decision-making structures(57), data safety monitoring boards (DSMB)(35,36), registries(58), and public-private parternships(19,20,22,23,28,33,51,54,59). Multidisciplinary collaboration across academia, government, and industry was emphasized(23). However, many of the existing research networks (e.g., HIV/AIDS, malaria, Zika, H1N1) were not mobilized for COVID-19 despite established infrastructure (24,40,57,60). This gap reflected systemic issues such as weak continuity, fragmented integration, and limited cross-regional coordination(61).

#### Infrastructure

Sites rapidly adopted decentralized and virtual workflows to sustain operations(48,54). Articles described new policies, procedures, and preparedness strategies, including repositories of trial templates and peer exchange on implementation practices(32,54,62,63). While acceleration enabled rapid study initiation, concerns about data quality persisted(64). Virtual visits, remote monitoring, and site reopening protocols illustrated how infrastructure evolved to maintain compliance and continuity(23–32,42,46).

#### Technology

Electronic consent presented early challenges during COVID-19, particularly in compliance and feasibility for patients in isolation(22,38,65,66). By the end of COVID-19, however, e-consent had become widely accepted.

Digital platforms supported remote enrollment, decentralized trial conduct, data capture, and virtual monitoring(23,39,46,59,67), in many instances modeled after telemedicine(68,69). Many sites navigated the best way to capture study data while maintaining fidelity of the protocol. The use of automated invitations, scheduled surveys, and technological innovations in remote visits and study design relieves site-stressors related to in-person visits while still allowing for fidelity to the protocol(39). Innovations expanded trial access to rural sites but were constrained by weak digital infrastructure and variable regulatory readiness(67,68).

#### Study Design

Duplicative and underpowered studies created operational strain and confusion(22,61,70). Conducting end-to-end digitalized protocols including describing the study setup in detail, evaluating its performance, and identifying points of success and failure was seen as necessary during COVID-19. When considering adaptations to study design during outbreaks, the use of pragmatic and adaptive study designs were commonly cited(23,39,69). Ethical tensions arose around informed consent and trial designs during crises, as Ebola patients’ critical condition and isolation complicated voluntary consent, while urgency to test experimental therapies drove adaptive and compassionate use trial designs challenging scientific rigor and long-term generalizability(32).

Some challenges in implementation and considerations of study design were related to managing limited healthcare resource (28,71), enrolling into multiple studies, and geographical distribution and engagement of sites(26,54,59). There were frequent calls for national-level coordination tools to prevent redundancy and prioritize research(33,46).

Drug repurposing was attempted widely but was often fragmented, demonstrating the need for coordinated prioritization and inclusion of stakeholders in study design. Allocating resources to incorporate research into medicine is one method to ensure appropriate outbreak response, but also to safeguard the future of clinical trials across the industry(28,38,41,46,47,50,65). By embedding research capacity directly into pandemic response, health systems can generate evidence in real time, thereby strengthening the credibility, infrastructure, and ethical standards of clinical trials. There is a need for collaboration among countries and manufacturers to define and develop frameworks to support sites, especially in resource constrained settings.

#### Communication

Remote staff training, monitoring, and site correspondence became standard during COVID-19. Similarly, from the site-manufacturer perspective, traditional site monitoring must be reconsidered during outbreaks. Thus, remote monitoring, which had not be used previously, was adopted during COVID-19. Additionally, literature described the support for remote data review and monitoring(33,35,54,55,72) under rapidly changes guidance(62,66).

Knowledge exchange and enhancing information sharing among clinical trial sites, was often accomplished with components of policy and community engagement(20,23,24,73). Expanding on the fundamental need for communication with participants via community engagement, a related component of this was instilling trust via transparency(23,32,63). Virtual workflows extended across trial operations(67), from screening and recruitment (67) to ongoing team coordination(54).

## Discussion

This review revealed persistent and complex operational challenges in conducting infectious disease clinical trials during pandemics. Misalignment between site processes and external systems—such as ethics and contracting bodies—delayed trial initiation, and in many cases, protocols were approved only after community infection rates had declined, undermining timeliness and feasibility. The review also noted a steady increase in reporting on clinical trial site operations over time, reflecting both an expanding evidence base and the growing relevance of this field.

This review consolidates global evidence on clinical trial operations during infectious disease pandemics and reveals an urgent need for more coordinated, collaborative, and transparent approaches. It is, to the author’s knowledge, the first scoping review to comprehensively examine site-level operations and adaptations in this context.

Decisions to pause non-COVID-19 research, prioritize COVID-19 studies, and restrict family access to patients significantly influenced site responses and disrupted studies within and beyond infectious disease domains(74). Simultaneously, multiple facilitators of success were identified, including scientific leadership, effective communication and data systems, protocol adaptation, early site engagement, streamlined regulatory processes, and the establishment of nested studies.

Articles consistently emphasized the need for multidisciplinary, trial-specific leadership with site representation from protocol development through implementation. Sites also require sponsor support to deploy digital tools, facilitate consent, manage data, and reduce protocol amendments. Adaptive and pragmatic trial designs were highlighted for their potential, though inconsistent application proved difficult for sites to implement(75) Several studies recommended avoiding poorly designed protocols(76,77), and encouraged aggregation of similarly designed trials into adaptive or nested formats(78–80).

Resource management challenges underscored the importance of stronger hospital-community research integration and outpatient infrastructure. Some institutions successfully redeployed staff (e.g., oncology personnel transitioning to COVID-19 studies(52), but persistent gaps in staff protection (e.g., vaccination) jeopardized operational continuity. Public-facing communication was also critical, with strategies such as simplifying scientific information, framing trials as care options (CRAACO), and embedding research in care pathways to build transparency and participant empowerment(81,82).

In resource-limited settings, centralized ethical review and sustained national funding were identified as vital for trial infrastructure(47,49,73). While direct-to-patient drug delivery and decentralized approaches were explored, they often introduced regulatory and logistical complexity. Inflated recruitment targets and weak feasibility assessments further strained operations, pointing to the need for more selective and realistic study design. Remote recruitment and advertising showed promise but remained among the most difficult operational components. Success in outpatient research depended on mobile teams, specialized staff, improved hospital-community coordination, and investments in technical infrastructure and training(83–85).

Global equity challenges were also evident. Preparedness requires strengthening infrastructure for infectious disease and critical care trials through multilateral cooperation among hospitals and institutions(76,77,86,87). Several articles noted disparities in authorship, with affiliations often not reflecting trial locations, highlighting long-standing inequities(45,69,88). Sites in low- and middle-income countries emphasized the importance of international collaboration through platforms that build local capacity(89). Sustained investment in pandemic preparedness, particularly for infectious disease and critical care trials, requires multilateral cooperation among hospitals and institutions(52,84,85,90).

This review had limitations. Database searches identified relevant material from disparate fields, and there were relatively few publications beyond those on COVID-19 which dominated the evidence base. The absence of risk-of-bias assessment or advanced synthesis prevented our ability to draw conclusions about the effectiveness of reported strategies(91). As a scoping review, no formal quality assessment of included records was performed(92). Replication could potentially yield a different set of included articles, though this is unlikely to alter the overall findings.

Notably, several included articles fell outside infectious disease but still offered useful reflections on operational adaptations during COVID-19. Comparing lessons across therapeutic areas may provide cross-cutting strategies for future emergencies. Further research is needed to identify which adaptations were most impactful, potentially informing a standardized triage checklist to guide site-level preparedness and decision-making.

### Conclusions

There is no single “gold standard” for conducting trials during pandemics. However, lessons from COVID-19 and other outbreaks underscore the importance of investing in site-level workforce development, integrating research into care pathways, and involving operational staff as core stakeholders. Industry and regulatory momentum is growing toward global preparedness frameworks and decentralized models, but further work is needed to make these approaches feasible at the site level.

At an industry level, there is growing momentum to develop global collaborative frameworks that clinical trial sites can leverage during pandemics and other emergencies. On the ground, manufacturers continue to explore and refine decentralized and hybrid models of trial conduct, though challenges persist in operationalizing these at sites. Tools such as operational checklists(93) offer promise as practical guides for implementation. While the FDA has provided guidance on decentralized research approaches, many questions remain, and such models, while promising, are not one-size-fits-all solutions.

These findings should inform both policy and practice. Permanent and flexible adaptations, such as operational checklists, integrated trial networks, and digital infrastructure, should be systematically incorporated into institutional and national planning exercises. A registry of institutional adaptations during pandemics and public health emergencies could support quality improvement and future readiness. Ultimately, advancing clinical research preparedness requires a holistic, systems-level approach that spans institutions, sectors, and borders.

## Data Availability

Data collected and analyzed can be made available upon request.

## Acknowledgements

The authors acknowledge Karen Robinson for her guidance during the planning and analysis stages of this review.

## Appendix

